# The detection and stability of the SARS-CoV-2 RNA biomarkers in wastewater influent in Helsinki, Finland

**DOI:** 10.1101/2020.11.18.20234039

**Authors:** Anna-Maria Hokajärvi, Annastiina Rytkönen, Ananda Tiwari, Ari Kauppinen, Sami Oikarinen, Kirsi-Maarit Lehto, Aino Kankaanpää, Teemu Gunnar, Haider Al-Hello, Soile Blomqvist, Ilkka T. Miettinen, Carita Savolainen-Kopra, Tarja Pitkänen

**Affiliations:** Finnish Institute for Health and Welfare, Expert Microbiology Unit, Kuopio, Finland; Tampere University, Faculty of Medicine and Health Technology, Tampere, Finland; Finnish Institute for Health and Welfare, Forensic Toxicology Unit, Helsinki, Finland; Finnish Institute for Health and Welfare, Expert Microbiology Unit, Helsinki, Finland; University of Helsinki, Faculty of Veterinary Medicine, Dept. Food Hygiene and Environmental Health, Finland

## Abstract

Wastewater-based surveillance of the severe acute respiratory syndrome coronavirus 2 (SARS-CoV-2) is used to monitor the population-level prevalence of the COVID-19 disease. In many cases, due to lockdowns or analytical delays, the analysis of wastewater samples might only be possible after prolonged storage. In this study, the effect of storage conditions on the RNA copy numbers of the SARS-CoV-2 virus in wastewater influent was studied and compared to the persistence of norovirus over time at 4°C, −20°C, and −75°C using the reverse-transcription quantitative PCR (RT-qPCR) assays E-Sarbeco, N2, and norovirus GII. For the first time in Finland, the presence of SARS-CoV-2 RNA was tested in 24 h composite influent wastewater samples collected from Viikinmäki wastewater treatment plant, Helsinki, Finland. The detected and quantified SARS-CoV-2 RNA copy numbers of the wastewater sample aliquots taken during 19–20 April 2020 and stored for 29, 64, and 84 days remained surprisingly stable. In the stored samples, the SARS betacoronavirus and SARS-CoV-2 copy numbers, but not the norovirus GII copy numbers, seemed slightly higher when analyzed from the pre-centrifuged pellet—that is, the particulate matter of the influent—as compared with the supernatant (i.e., water fraction) used for ultrafiltration, although the difference was not statistically significant. Furthermore, when wastewater was spiked with SARS-CoV-2, linear decay at 4°C was observed on the first 28 days, while no decay was visible within 58 days at −20°C or −75°C. In conclusion, freezing temperatures should be used for storage when immediate SARS-CoV-2 RNA analysis from the wastewater influent is not possible. Analysis of the particulate matter of the sample, in addition to the water fraction, can improve the detection frequency.

## 1 Introduction

Wastewater-based surveillance has been proposed for a rapid and resource-efficient way to monitor the population-level prevalence of severe acute respiratory syndrome coronavirus 2 (SARS-CoV-2), an etiological agent of the current global pandemic of coronavirus disease 2019 (COVID-19) (WHO, 2020; Ahmed *et al*., 2020a; Lodder and de Roda Husman, 2020; Mallapaty, 2020; Medema *et al*., 2020a; Orive *et al*., 2020). The wastewater-based approach can provide an effective way for identifying community-level hotspots of the infection and has been proposed as an early warning tool, providing extra time for actions to suppress the spread of infections before the availability of clinical surveillance information (La Rosa *et al*., 2021; Kitajima *et al*., 2020; Thompson *et al*., 2020). During the COVID-19 pandemic, the analytical capacities to study the SARS-CoV-2 RNA biomarkers from wastewater samples has been limited due to the laboratory and campus lockdowns at many locations with remote work recommendations. Furthermore, the lack of required methodological resources and validated methods, and delays in the deliveries of laboratory reagents and supplies (like the delayed availability of the plasticware required for ultrafiltration) due to global shortage, have hampered the immediate analysis of the SARS-CoV-2 RNA from the collected wastewater samples. Subsequently, on many occasions, researchers aiming to contribute to the fight against COVID-19 have been forced to store the collected samples and wait for the analytical resources to become available.

It has been claimed that the SARS-CoV-2 might have already circulated in many locations before the first reports from Wuhan, China, were received (Deslandes *et al*., 2020; La Rosa *et al*., 2021). Previously stored samples—like those of wastewater influents collected during the worldwide poliovirus monitoring (La Rosa *et al*., 2021; Hovi *et al*., 2012), together with stored respiratory samples from patients (Deslandes *et al*., 2020)—need to be analyzed to trace back to the first introduction of SARS-CoV-2 in different cities of the world. After the summer months of 2020, where the COVID-19 pandemic had a very slow or stagnant phase in countries like Finland, the resources for the quantitation of SARS-CoV-2 RNA from wastewater became more readily available. The analysis of stored samples might finally be possible in many locations, although not all bottlenecks to analysis availability have been tackled completely and the immediate actions required to tackle the next waves of the COVID-19 pandemic might again slow down the analysis of the previously stored samples.

It is important to determine the effect of storage conditions on the reliability of the SARS-CoV-2 RNA results as there is usually always a time lag between the sampling and the start of the laboratory examination due to the transportation and other logistical arrangements (Medema *et al*., 2020a). So far, the persistence of SARS-CoV-2 RNA in stored wastewater samples has only been studied at higher temperatures, 4°C, 15°C, 25°C, and 37°C (Ahmed *et al*., 2020b) and at 20°C, 50°C, and 70°C (Bivins *et al*., 2020). Furthermore, some stability information is obtained from other coronaviruses, like human coronavirus 229E (Gundy *et al*., 2008) and SARS-CoV-1 (Wang *et al*., 2005), or from surrogate organisms, such as murine hepatitis virus (MHV) (Casanova *et al*., 2009; Ye *et al*., 2016). This study aimed to establish the SARS-CoV-2 RNA stability characteristics in cold and freezing temperatures in order to evaluate the usability and reliability of the SARS-CoV-2 RNA data gathered from previously collected and stored wastewater samples. To do this, we followed the decay of the SARS betacoronavirus envelope gene by using an E-Sarbeco RT-qPCR assay (Corman *et al*., 2020) and the decay of the SARS-CoV-2 nucleocapsid gene by using an N2 assay (Lu *et al*., 2020) in wastewater influent aliquots stored at 4°C, −20°C, and −75°C over time.

As a reference virus, the decay of norovirus GII, a non-enveloped human enteric virus known for its persistence in the environment (Kauppinen and Miettinen, 2017) and commonly found in municipal wastewater, was used. Further, this study has also compared the detection of SARS-CoV-2 RNA copy numbers from the pre-centrifuged pellets—that is, the particulate matter of the influent—with the supernatant (i.e., water fraction) used for ultrafiltration. This is the first study reporting the testing and detection of SARS-CoV-2 RNA from municipal wastewater in Finland.

## 2 Materials and Methods

### 2.1 Wastewater influent sampling and storage

Table 1 shows the characteristics of the wastewater influent samples collected from Viikinmäki wastewater treatment plant (WWTP), Helsinki, Finland.

**Table 1.** The characteristics of the wastewater influent at Viikinmäki WWTP, Helsinki, Finland. The data obtained from the WWTP and the national infectious disease register.

| Sampling time | From 19.4.2020 07:00<br>to 20.4.2020 07:00 | From 24.5.2020 07:00<br>to 25.5.2020 07:00 |
| --- | --- | --- |
| COVID-19 cases reported in the sewerage network area within the 14 days preceding the sampling event | 1,076 | 409 |
| Total inflow during sampling (m<sup>3</sup>/d) | 306,019 | 251,741 |
| Composite sampling interval (m<sup>3</sup>) | 18.8 | 18.8 |
| Influent temperature, °C (mean; min–max) | 12.0; 10.4–13.2 | 13.3; 12.4–14.2 |
| pH | 7.4 | 7.4 |
| BOD (mg/l) | 205 | 247 |
| COD (mg/l) | 404 | 541 |
| TSS (mg/l) | 204 | 302 |
| N<sub>tot</sub> (mg/l) | 42 | 54 |
| P<sub>tot</sub> (mg/l) | 5.1 | 6.4 |
| NH<sub>4</sub>-N (mg/l) | 32 | 35 |

Abbreviations: na: not available; BOD: biochemical oxygen demand; COD: chemical oxygen demand; TSS: total suspended solids; N<sub>tot</sub>: total nitrogen concentration; P<sub>tot</sub>: total phosphorus concentration; NH<sub>4</sub>-N: ammonium nitrate concentration.

The Viikinmäki WWTP serves the population of 860,000 inhabitants from the municipalities of Helsinki, Sipoo, Kerava, Tuusula, Järvenpää, Pornainen, Mäntsälä, and Vantaa. Samples were collected from the influent before any treatment following the national guidance of biosafety precautions for WWTP personnel. Subsamples of 5,000 mL and 2,000 mL of 24-hour composite wastewater influent were collected from 6°C refrigerated 20,000 mL sample container during 19–20 April 2020 and 24–25 May 2020, respectively, and transported to the laboratory in cool boxes within 24–26 hours. Upon arrival to the laboratory, the samples were divided into 30 ml aliquots and stored at temperatures of 4°C, −20°C, and −75°C for later analysis.

### 2.2 The ultrafiltration method for the detection of SARS-CoV-2 RNA in the stored samples

For testing the presence and stability of SARS-CoV-2 RNA, the influent sample taken during 19–20 April 2020 was used. Frozen sample aliquots were thawed overnight at a temperature of 4°C before analyses.

Prior to ultrafiltration performed after storage of samples for 29, 64, and 84 days, two 30 ml aliquots were combined and larger particles were removed from triplicate 60 ml influent samples with pre-centrifugation at 4,654 g for 30 min without brake as previously described (Medema *et al*., 2020b). Then, the supernatants obtained (57–59 ml) were concentrated using Centricon Plus-70 centrifugal filters (a 10K device, UFC701008, Millipore Corporation). Before use, the ultrafilters were washed with sterile deionized water, following the manufacturer’s instructions. The ultrafiltration was conducted at 3,000 g for 25 min, followed by concentrate collection with 1000 g for 2 min, producing approximately 400 µL of concentrate.

In addition to the filtrated supernatant, the pellets from the suspended solids removal step (pre-centrifugation) were included into analysis soon after the importance of considering both liquid and solid fractions of wastewater samples when analyzing enveloped viruses such as SARS-CoV-2 had been realized (see, e.g., Ahmed *et al*., 2020c). The pellets were re-suspended using widened pipette tip with approximately 500 µL of supernatant and analyzed from the samples after 84 days of storage. The concentrate volume was normalized to 700 µL for all the filtrated supernatants and pre-centrifugation pellet samples by suspending them in the filtrate and supernatant, respectively.

All treatment steps were carried out at room temperature. Immediate nucleic acid extraction was conducted using 300 µL of the final concentrated sample while the rest of the concentrate was preserved at −75°C for later use.

The detection and quantitation of the viral targets entailed the determination of the copy numbers of SARS betacoronavirus with an E-Sarbeco assay (Corman *et al*., 2020) and of SARS-CoV-2 with an N2 assay (Medema *et al*., 2020b; Lu *et al*., 2020). Also, the copy numbers of norovirus GII (Kauppinen and Miettinen 2017) were analyzed.

### 2.3 The decay of SARS-CoV-2 RNA in wastewater stored at cold temperatures

The decay characteristics of SARS-CoV-2 RNA were studied at the time points of 0, 3, 7, 10, 14, and 21 days after the start of the experiment. Additionally, the effect of long-term storage was tested at 4°C after 28 days and at −20°C and −75 °C after 58 days. To reach sufficiently high copy numbers for the experiment, the influent sample taken during 24–25 May 2020 was spiked with a 1:1,000 dilution of SARS-CoV-2 inoculum prepared from the nasopharyngeal swab of a COVID-19 patient. The produced spiked wastewater influent was divided into 57 microcentrifuge tubes in portions of 300 µl. Triplicate tubes were stored in the dark at temperatures of 4°C, −20°C, and −75°C and taken to direct nucleic acid extraction and subsequent reverse transcriptase-quantitative PCR (RT-qPCR) analysis. Frozen samples were thawed on ice before the analysis. The copy numbers of SARS betacoronavirus RNA were quantified with an E-Sarbeco assay. The specificity of the E-Sarbeco assay was confirmed by Sanger sequencing of the selected PCR products. The presence and quantitation of SARS-CoV-2 RNA in the sample material was confirmed using the N2 assay.

### 2.4 Nucleic acid extraction and the RT-qPCR of the viral targets

For nucleic acid extraction, a Chemagic Viral300 DNA/RNA extraction kit was used with a Chemagic-360D semi-automated nucleic acid extraction instrument (Perkin-Elmer, USA). For inactivation, 300 µl of Lysis Buffer 1 and 300 µl of the sample (concentrate or a re-suspended pre-centrifugation pellet) were mixed and incubated in room temperature for at least 10 minutes. Poly(A) RNA and Proteinase K were added into the lysates and the extraction protocol was executed according to the manufacturer’s instructions. Prior to downstream analysis, an additional purification of the nucleic acids extracted from the pre-centrifugation pellets was done using a OneStep™ PCR Inhibitor Removal Kit (Zymo Research, USA) according to the manufacturer’s instructions.

The RT-qPCR assays were performed using a QuantStudio 6 Flex real-time PCR system (Applied Biosystems, ThermoFisher Scientific). For SARS betacoronavirus quantitation with an E-Sarbeco assay targeting envelope protein (Corman *et al*., 2020), RT-qPCR reactions were carried out in a final volume of 25 μl, containing a 5 μl RNA sample, 0.4 μM primers, a 0.2 µM probe, and 6.25 μl TaqMan Fast Virus 1-step Master Mix (Applied Biosystems, ThermoFisher Scientific). For SARS-CoV-2, an N2 assay targeting nucleocapsid protein (Medema *et al*., 2020b; Lu *et al*., 2020) was conducted in the same manner except that both primers and probe were used in a final concentration of 0.2 µM. For both assays, the thermal cycling conditions were as follows: 50°C for 5 min for reverse transcription, followed by 95°C for 20 s, and then 45 cycles of 95°C for 15 s and 58°C for 1 min. All the reactions considered positive exhibited Ct value below 40. The copy numbers of norovirus GII were quantified from the samples using RT-qPCR, as previously described (Kauppinen and Miettinen, 2017).

For quantitation of the RT-qPCR targets, standard curves for each run were generated by using serial dilutions of control materials with a known quantity. In the E-Sarbeco assay and N2 assay, control plasmids pEX-A128-nCoV_E_Sarbeco (Eurofins, Luxembourg) and 2019_nCoV_N Positive Control (IDT, USA) were used, respectively. Before use, the pEX-A128-nCoV_E_Sarbeco plasmid (100 ng µl^-1^) was digested with FastDigest NotI (ThermoScientific, USA) and purified using Illustra GFX PCR DNA and a Gel Band Purification Kit (GE Healthcare, UK) according to the manufacturer’s instructions. For the N2 assay, the plasmid was used without linearization following the manufacturer’s instructions. The quantitation of the norovirus GII assay was done using an artificial gene fragment (gBlocks, IDT).

### 2.5 Confirmation of the SARS-CoV-2 target by sequencing

The selected PCR products from E-Sarbeco assay were run by gel electrophoresis and approximately 113 bp long bands were cut from the gel. The purification was done using a QIAquick Gel Extraction Kit (Qiagen, Germany) and, further, the sequencing was done in both directions by ABI BigDye™ v3.1 Chemistry (Applied Biosystems, USA) according to the manufacturer’s protocol. The sequences were verified and aligned using Geneious 11.1.3 (Biomatters Ltd, New Zealand). The pair-end gene sequence of the E gene PCR product generated from a spiked wastewater sample was: ACTTCTTTTTCTTGCTTTCGTGGTATTCTTGCTAGTTACACTAGCCATCCTTACTGCGCTTCGAT. The consensus sequence was identified using BLAST analysis (https://blast.ncbi.nlm.nih.gov/Blast.cgi).

### 2.6 Safety measures, quality assurance, and controls

To ensure the safety of the laboratory work, wastewater and patient samples were processed as pair-work in a level 2 biosafety cabinet using cuffs, double gloves, and a laboratory coat. Only airtight sealed tubes were handled outside the biosafety cabinet. Seventy per cent ethanol was sprinkled on the outer surfaces of the tubes and airtight rotor lids and also used during the opening of the centrifuge lid after a one- to two-minute waiting time following the centrifugation in order to prevent possible aerosols from entering into the air. To estimate the recovery efficiency of the ultrafiltration method, an enveloped virus control similar to SARS-CoV-2 was not available. Instead, a non-enveloped mengovirus was used as an internal process control following the principles of technical specification ISO/TS 15216-1:2013. Spiked mengovirus strain VMC0 (ATCC VR-1597, CeeramTOOLS, France) was used.

Each RNA extraction set included reagent blanks (300 µl PBS or sterile deionized water) and a SARS-CoV-2 positive process control (300 µl of 1:500 diluted nasopharyngeal swab sample from a COVID-19 positive patient, dissolved into PBS and inactivated at 60oC for 90 min). In RT-qPCR, standard curves and no template controls were included in each run. The actual wastewater volume analyzed for each PCR assay (mL) was 3.8–4.0 mL for the authentic samples analyzed using the Centricon Plus-70 concentration method and 0.07 mL in the inoculation experiment with direct extraction. Where possible, extracted RNA samples and their ten-fold dilutions were analyzed in duplicate. Ten-fold diluted samples were used to estimate the presence of inhibition: inhibition was considered significant if the gene copy number detected from the diluted sample was two times higher than the gene copy number detected from the undiluted sample. When applicable, the average results from both the diluted and undiluted sample were used to generate the final data.

### 2.7 Statistics

The viral RNA copy numbers were log-transformed into log10 before statistical analysis. The standard error with a 95% confidence interval was calculated with the following formula:

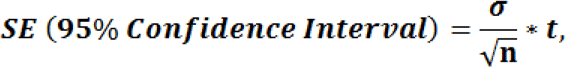

where σ is a standard deviation, n is the number of samples, and t is a critical value at the probability level of 0.025 (two-tailed) from a t-distribution table at (n-1) degrees of freedom.

The differences between the copy numbers in samples stored at the different temperatures were compared with the Kruskal-Wallis test followed with the Dunn post hoc test. The overlapping upper and lower confidence interval of mean copy numbers from E-Sarbeco and N2 assays generated from pellet and supernatant fractions were compared, as previously described (Tiwari *et al*., 2019).

The decay characteristics at the storage temperatures were analyzed with the GInaFiT Version 1.7 (Geeraerd and Van Impe Inactivation Model Fitting Tool) freeware add-in for Microsoft® Office 365® Excel (Geeraerd *et al*., 2005), as done earlier (Tiwari *et al*., 2019, Han *et al*., 2020). GInaFiT is a commonly used tool for modeling the decay of microbes or their genetic markers. It tests nine different types of decay model, such as log-linear and biphasic models. In the GInaFiT tool, the regression analysis was performed (log_10_(N_t_) vs. time) for each viral target including the prediction of times needed for 90% and 99% reduction at the storage temperatures in the wastewater influent, as previously described by Casanova *et al*. (2009). Statistical analysis was done using IBM SPSS statistics.

## 3 Results

The SARS betacoronavirus, SARS-CoV-2, and norovirus GII RNA were detected from the wastewater sample collected during 19–20 April 2020 from the Viikinmäki WWTP, Helsinki, Finland. The quantity of SARS betacoronavirus and SARS-CoV-2 RNA when stored at 4°C, −20°C, and −75°C and analyzed with E-Sarbeco and N2 assays, respectively, remained stable at all storage temperatures for up to 84 days (Table 2). At the same time, norovirus GII RNA copy numbers reduced about 1-log_10_ during the time interval between 29 and 84 days of storage (Table 2). The copy numbers of SARS betacoronavirus and SARS-CoV-2 RNA were slightly more frequently quantified with a higher total mean from the pellet fraction of the samples stored for 84 days, that is, the particulate matter from the pre-centrifugation step, rather than from ultrafiltrated supernatant fraction (Table 2, E-Sarbeco and N2 assays). At the same time, with a non-enveloped virus type, norovirus GII, used as a different reference to enveloped SARS and SARS-CoV-2 viruses, the target mean difference in the quantified copy numbers between the pellet and supernatant fractions was the opposite.

**Table 2.**
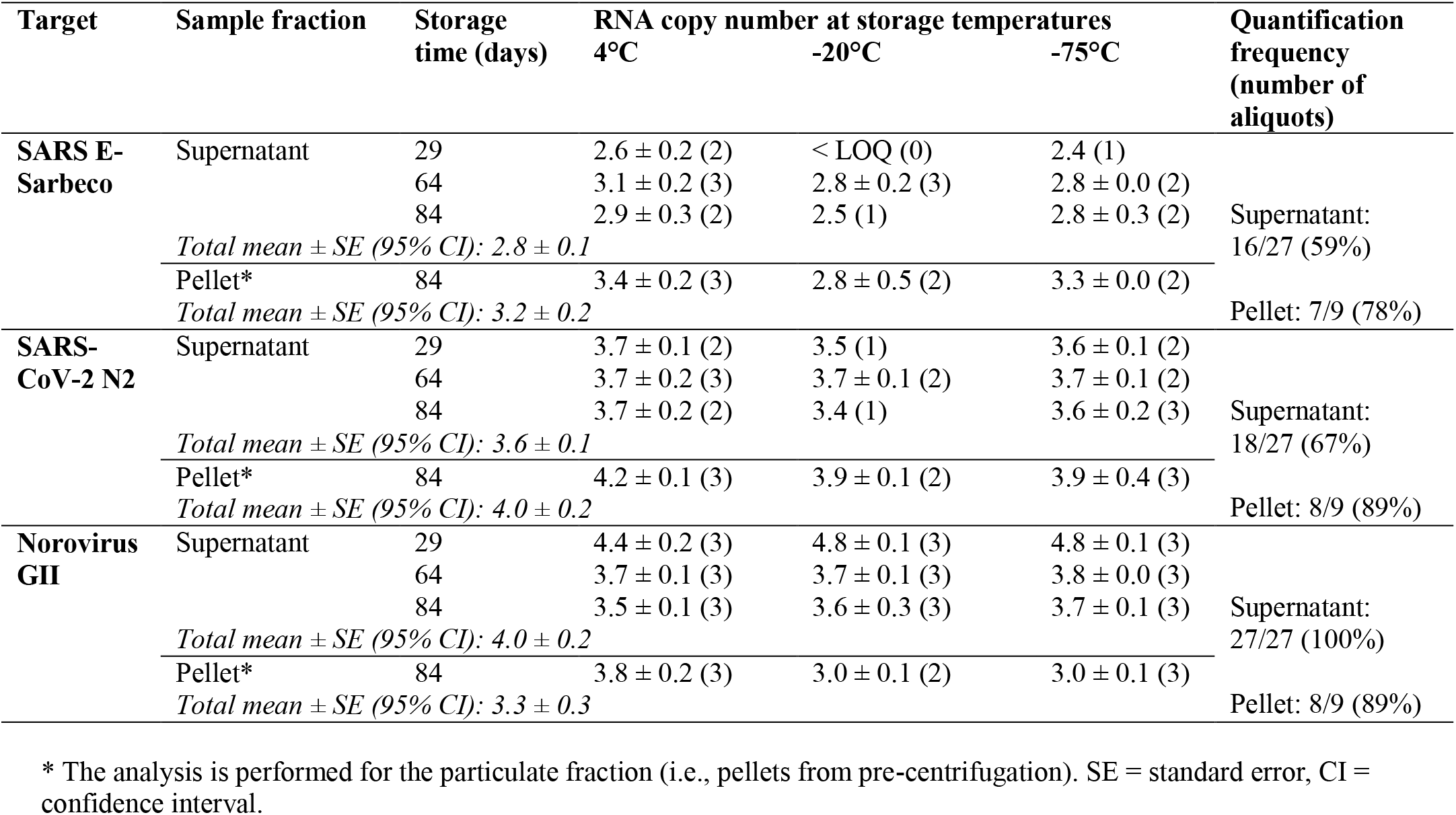
The mean copy numbers of virus RNA markers (mean ± standard deviation, log_10_ copies 100 ml^-1^) detected with SARS betacoronavirus E-Sarbeco, SARS-CoV-2 N2 and norovirus GII assays in Viikinmäki WWTP influent wastewater sample taken 19–20 April 2020 and stored over 84 days at 4°C, −20°C, and −75°C. The analysis at each time and temperature point was performed from triplicate aliquots. The number of replicates with results above the quantification limit (LOQ) are shown in parenthesis.

The copy numbers from days 0–28 remained unknown for the sample taken 19–20 April 2020 (Table 2) since during that time the method for the immediate analysis of the viral biomarkers from influent samples was not available. Therefore, a Viikinmäki WWTP influent wastewater sample taken five weeks later, during 24–25 May 2020, was used to experimentally characterize the decay rate of SARS-CoV-2 RNA copy numbers during the first month of the cold storage of wastewater. Before spiking, the sample contained the target RNA of the levels 3.1 and 3.8 log_10_ copies 100 ml^-1^ when analyzed using E-Sarbeco and N2 assays, respectively. After a spike with a nasopharyngeal swab from a COVID-19 patient, the copy numbers of the RNA targets in the beginning of the experiment reached the level of 7.4 and 8.1 log_10_ copies 100 ml^-1^ in E-Sarbeco and N2 assays, respectively.

When the E-Sarbeco assay was used, no statistically significant difference between the storage temperatures was noted (Figure 1, *p* = 0.258, Kruskal-Wallis), although after two weeks of storage and thereafter, the sample aliquots stored at freezing temperatures (−20°C and −75°C) seemed to produce higher quantities than refrigeration (4°C). When the N2 assay was used, a statistically significant difference between the storage temperatures was observed (Figure 1, *p* = 0.039, Kruskal-Wallis). A linear decay of SARS-CoV-2 RNA was observed at 4°C while no decay was visible within 58 days at the freezing temperatures of −20°C and −75°C (Figure 1). Regression analysis, calculated with GInaFiT, was done for the sample aliquots stored at 4°C (Table 3). The decay was more linear and slightly faster when N2 was used as a target (*R*^2^ = 0.99) when compared to E-Sarbeco assay results (*R*^2^ = 0.59).

**Table 3.** The decay characteristics of the SARS-CoV-2 spike (log_10_ copies 100 ml^-1^) in wastewater influent at 4°C, enumerated with E-Sarbeco and N2 assays. Samples stored at −20°C and −75°C were not suitable for decay modeling as no decay was observed.

| Assay | Decay Rate (k) | $R^2$ | Square Difference | $T_{50}$ (days) | $T_{90}$ (days) | $T_{99}$ (days) |
| --- | --- | --- | --- | --- | --- | --- |
| E-Sarbeco | 0.04 ± 0.2 | 0.59 | 0.15 | 16 | 52 | 105 |
| N2 | 0.06 ± 0.0 | 0.99 | 0.00 | 11 | 36 | 73 |

**Figure 1.**
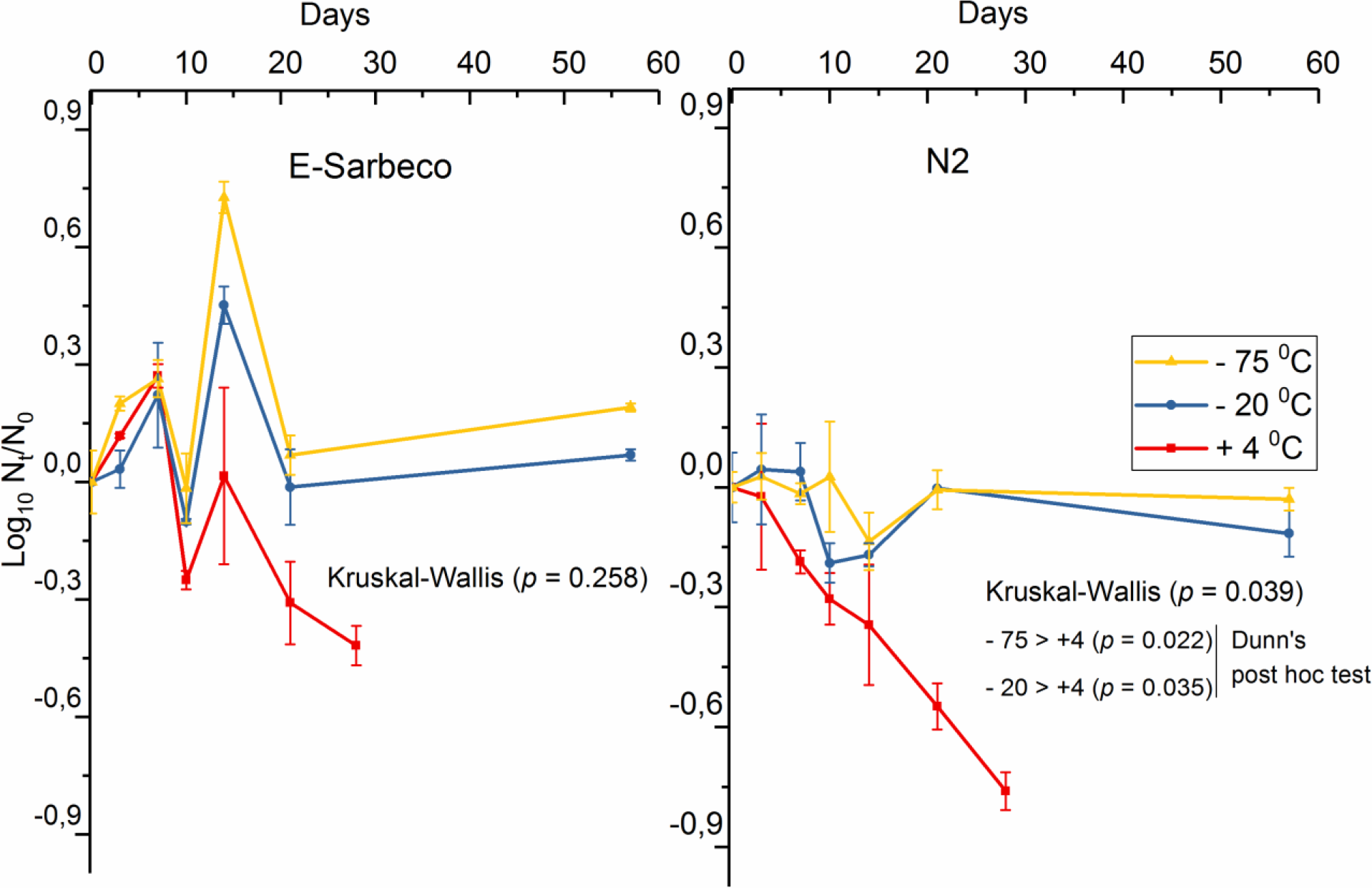
The decay curve of SARS-CoV-2 spike (log_10_ copies 100 ml^**-1**^) in wastewater influent at 4°C, −20°C, and − 75°C, enumerated with E-Sarbeco and N2 RT-qPCR assays.

## 4 Discussion

This is the first study reporting the detection of SARS-CoV-2 RNA in wastewater influent in Finland. The presence of the biomarkers of SARS-CoV-2 in wastewater influent samples collected during April and May 2020 from Viikinmäki WWTP, Helsinki, Finland, is in agreement with the national infectious disease register data regarding the confirmed COVID-19 cases in the municipalities of the Viikinmäki WWTP sewerage network area preceding the sampling and with studies conducted earlier in other countries (e.g., Ahmed *et al*., 2020a; Haramoto *et al*., 2020; La Rosa *et al*., 2020a; Medema *et al*., 2020b; Mlejnkova *et al*., 2020; Arora *et al*., 2020). In the sample aliquots stored at 4°C, −20°C, and −75°C and analyzed after storage herein, the SARS-CoV-2 RNA seemed surprisingly stable in these cold storage temperatures from 29 days to 64 and 84 days prior to carrying out the sample concentration and nucleic acid extraction. Further, the results generated by spiking the wastewater influent matrix with a diluted nasopharyngeal swab sample from a COVID-19 patient and following the RNA signal from experiment initiation (day 0) to 28 and 58 days in refrigerated and freezing conditions, respectively, confirmed the stability of SARS-CoV-2 RNA, especially when stored in freezing temperatures.

A limitation of the study set-up was that we were not able to follow SARS-CoV-2 copy numbers during the first 28 days of storage using the real wastewater samples. Instead, we conducted a separate spike-in study to reveal the decay characteristics between 0 and 28 days. The conventional route of transmission of SARS betacoronaviruses is via respiratory droplets passing through the nasopharynx. A nasopharyngeal swab sample was used herein in order to gain high enough quantities of SARS-CoV-2 for the spike-in experiment. However, the survival kinetics of SARS-CoV-2 after passage through the gastrointestinal tract and, furthermore, after the transport of the viral particles—first in the sewerage system to the WWTP and then in a sample container to the laboratory—might be different than those of a highly contagious nasopharyngeal sample. While the finding of viral RNA in stools or wastewater does not imply that the virus is viable and infectious (Foladori *et al*., 2020), the two experiments carried out herein (the analysis of actual wastewater samples after storage and the spike-in study) both indicated the stability of the RNA signal during storage in freezing temperatures. Therefore, this study has assumed the decay characteristics of the RNA from fresh virus particles from COVID-19 patient may not differ from virus RNA from wastewater samples. The wastewater system represents virus particles that have ended up there from the fecal materials, urine, coughing, sneezing, and sputum of infected people, with the majority of virus particles possibly originating from feces (WHO, 2020).

While the viruses previously analyzed by environmental microbiologists are most commonly non-enveloped viruses—such as poliovirus, norovirus, adenovirus, or enterovirus—SARS-CoV-2 is an enveloped virus. Currently, the protocols for SARS-CoV-2 testing from wastewater samples vary a lot despite the multiple efforts to harmonize the virus concentration and quantitation approaches (Hill *et al*., 2020; Kitajima *et al*., 2020; Kordatou *et al*., 2020). As the aim is to detect the virus at low concentrations in wastewater, the efficiency of the procedures employed for sampling, preserving, and processing samples are critical (Medema *et al*., 2020a) and there is a need for developing standardized and reliable virus quantification protocols (Orive *et al*., 2020).

Interestingly, while the SARS-CoV-2 RNA remained almost stable at 4°C, −20°C, and −75°C when the wastewater influent was stored up to 84 days, norovirus reduced by about 1 log_10_ value during the same time. This observation indicates that (actually against the current belief; Hill *et al*., 2020) the persistency of non-enveloped viruses is not necessarily higher than the persistence of enveloped viruses in cold environmental conditions. Based on previous reports, it is known that coronaviruses decline rapidly at ambient temperatures in wastewater (Ahmed *et al*., 2020b; Gundy *et al*., 2008; La Rosa *et al*., 2020b; Silverman and Boehm, 2020). Wang *et al*. (2005) recovered SARS-CoV-1 after at least 14 days in wastewater stored at 4°C while they reported it surviving only two days at 20°C. Ye *et al*. (2016) concluded that, with MHV surrogate, the inactivation kinetics of the enveloped viruses are significantly slower in wastewater at 10°C compared to 25°C. Previously, with surrogate coronaviruses, transmissible gastroenteritis, and MHV, Casanova *et al*. (2009) concluded that at 4°C there was a <1 log_10_ infectivity decrease for both viruses after four weeks. More recently, Ahmed *et al*. (2020b) reported that, by using an N1 gene target for SARS-CoV-2 RNA in untreated wastewater, the mean first-order decay rate constant (*k*) for SARS-CoV-2 was 0.084/day at 4°C. In the present study, SARS-CoV-2 RNA copy numbers exhibited a linear decay with a *k* value of 0.04 and 0.06 for N2 and E-Sarbeco gene targets in wastewater, respectively, over 21 days when stored at 4°C. On the contrary, the RNA count in Viikinmäki WWTP influent remained unchanged over 58 days in the wastewater influent stored at −20°C and − 75°C. Overall, the inactivation of coronaviruses in the wastewater seems highly dependent on temperature. However, between the samples and sampling locations, the level of organic matter, and the presence of other microbes could affect the persistence.

Of the two biomarker assays for SARS-CoV-2 employed herein, the SARS-CoV-2 RNA quantified using the N2 assay exhibited a slightly higher decay rate than the SARS betacoronavirus RNA copy numbers obtained using the E-Sarbeco assay. The slightly more rapid decay of the N2 assay target compared with the E-Sarbeco assay target may indicate the differential decay rates of nucleocapsid and envelope genes (Mousavizadeh and Ghasemi, 2020). More information is needed on the stability of different marker genes used for wastewater-based SARS-CoV-2 surveillance.

Overall, the information about the fate of enveloped viruses in municipal wastewater has been scarce. When Ye *et al*. (2016) studied the survival and partitioning behavior of model viruses in raw wastewater samples, up to 26% of the enveloped MHV and *Pseudomonas* phage ϕ6 viruses adsorbed to the solid fraction of wastewater compared with 6% of the non-enveloped bacteriophages (MS2 and T3). Although more research is needed, SARS-CoV-2 in WWTP influent could also be associated with the particulate fraction of the sample.

It is noteworthy that the ultrafiltration method commonly used for wastewater SARS-CoV-2 RNA analysis includes a pre-centrifugation step, wherein part of the particulate fraction is removed from the analysis (Medema *et al*., 2020b). Such an approach might not be optimal for enveloped viruses such as SARS-CoV-2, as was recently shown in a MHV surrogate study where concentration methods using both liquid and solid fractions of wastewater produced the best yields (Ahmed *et al*., 2020c). When we used the ultrafiltration method for the Viikinmäki WWTP samples stored for 84 days at 4°C, −20°C, and −75°C, the quantification frequency and the quantities of SARS-CoV-2 RNA biomarkers were slightly higher when analyzed from pre-centrifuged pellets rather than from the water fraction without the particulate matter. From the same samples, the quantification frequency and the copy numbers of the norovirus RNA were on average higher when analyzed from supernatants when compared with pellets. The finding may indicate the higher affinity of an enveloped virus like SARS-CoV-2 to attach onto the particulate matter of wastewater when compared with non-enveloped viruses like norovirus. Indeed, instead of only studying wastewater influent, research groups have recently reported SARS-CoV-2 RNA detection in sewage sludge, proposing that the sludge could serve as a better indicator for COVID-19 outbreak dynamics than WWTP influent (Kocamemi *et al*., 2020; Peccia *et al*., 2020).

## 5 Conclusions

- SARS-CoV-2 RNA was detected for the first time in the 24 h composite influent wastewater sample collected from Viikinmäki WWTP, Helsinki, Finland, during 19–20 April 2020.
- The SARS-CoV-2 RNA remained detectable during storage periods of 29, 64, and 84 days at 4°C, −20°C, and −75°C when analyzed using E-Sarbeco and N2 RT-qPCR assays.
- The SARS-CoV-2 RNA quantification frequency and copy numbers were slightly higher when the particulate matter of the influent was analyzed, compared with the pre-centrifuged supernatant fraction used for ultrafiltration.
- In the sample pre-treatment procedures for ultrafiltration, the pellet from the initial larger particle removal step should not be discarded when the samples are analyzed after long-term storage. In the analysis of fresh wastewater samples, the necessity to also analyze the pellet should be further investigated.
- A linear decay of spiked SARS-CoV-2 RNA was observed at 4°C while no decay was visible within 58 days at the freezing temperatures. In cases when immediate SARS-CoV-2 RNA analysis from the wastewater influent is not possible, storage at −20°C or −75°C should be preferred.

## Data Availability

Data available within the article.

## Acknowledgements

The authors would like to express special thanks to the personnel of Helsinki Region Environmental Services Authority HSY and Director Mari Heinonen at the HSY Wastewater Services Department for their support during the project’s establishment and WWTP sampling arrangements. The personnel of the Health Security Department at the Finnish Institute for Health and Welfare are acknowledged with special thanks given to Oskari Luomala and Riikka Airaksinen for their help with the communicable disease statistics and to Tiina Heiskanen, Marjo Tiittanen, Tarja Rahkonen and Arja Moilanen for their assistance in the laboratory. This study was supported by Finnish Government supplementary budget for Covid-19 research and the Ministry of Agriculture and Forestry (grant number 4400T-0807).

